# CovidSIMVL – Agent-Based Modeling of Localized Transmission within a Heterogeneous Array of Locations – Motivation, Configuration and Calibration

**DOI:** 10.1101/2020.11.01.20217943

**Authors:** Kenneth A. Moselle, Ernie Chang

## Abstract

CovidSIMVL is an agent-based infectious disease modeling tool that is designed specifically to simulate localized spread of infectious disease. It is intended to support tactical decision-making around localized/staged re-institution of pre-pandemic levels and patterns of social/economic/health service delivery activity, following an initial stage of pan-societal closures of social/economic institutions and broad-based reductions in services.

By design, CovidSIMVL supports the generation of dynamic models that reflect heterogeneity within and between a network of interacting localized contexts. This heterogeneity is embodied in a hierarchically organized set of rules. Primary rules reflect the pathophysiology of transmission. Secondary rules (“HazardRadius” and “Mingle Factor” in CovidSIMVL) relate transmission to proximity and movement within physically demarcated and relatively contained spaces (“Universes”). Tertiary rules (“Schedules”) relate probabilities of transmission to movement of people between a network of localized contexts (a CovidSIMVL “Multiverse”).

This report focuses mainly on calibration of secondary rules. To calibrate the HazardRadius and MingleFactor parameters, growth curves were generated with CovidSIMVL by setting different configurations of values on those two proximal determinants of viral transmission. These were compared to the characteristic shapes of curves generated by equation-based compartmental models (e.g., SEIR models) that fit different real-world datasets embodying different reproduction numbers (R_0_).

By operating with parameter values in CovidSIMVL that generate “real-world” growth curves, the tool can be used to produce plausible simulations of localized chains of transmission. These include transmission among different groups of persons (e.g., staff, patients) who are co-located within a single setting such as a long-term care facility. The Multiverse version of CovidSIMVL can be used to simulate localized cross-over transmission among arrays consisting of both unaffected and impacted contexts and associated sub-populations, *via* agents who interact within and across arrays of contexts such as schools, multigenerational families, recreational facilities, places of work, emergency shelters for homeless persons, or other settings in which people are in close physical proximity.

## INTRODUCTION

### Context – why/when are agent-based models fit for purpose?

Ogden^1^ reviews two streams of activity within the Public Health Agency of Canada (PHAC) to assess the impact of different non-pharmaceutical interventions (NPIs) on spread of SARS-CoV-2. These two approaches, which Ogden and associates characterize as complementary, consist of equation-based compartmental models, and agent-based models.

The compartmental equation-based approaches typically operate on the basis of a “mass action incidence” model that assumes all individuals within a population make contact with one another at an identical rate, and have identical probabilities of transmission to any other member of the population per unit time. This assumption may be reasonable when the number of infective persons in a population is large and cases are geographically well-distributed.^2,3^ To introduce some measure of sub-population-level heterogeneity, meta-population variants of equation-based compartmental models, including “patch” variants,^4^ may capture some of the structural inhomogeneity in contact patterns that reflect “real-world” features, e.g., children in schools. However, use of compartmental models to inject local realism into models of infectious disease transmission becomes challenging at the level of computation and estimation, when the number of units (patches) associated with subsets of observations grows in order to incorporate even a coarse level of local topographical realism into models, and when the network of connections between patches grows in order to capture the dynamics of movement associated with non-localized secondary spread of the infection.^5^

Compartmental equation-based models describe/predict rates of change over time and are well-suited to the task of motivating policy/regulatory bodies to institute large-scale protections necessary to mitigate risks, when the conditions necessary for continued spread are well distributed throughout the population. They are particularly relevant when reproduction numbers are relatively high (e.g., when rates are increasing exponentially) at the same time that the infection is distributed in a manner that crosses multiple population/clinical demographics and geographical boundaries.

However, Brauer, as well as Ball et al.^6^ note that compartmental models are challenged to capture the network structures that govern transmission in the early stages of a pandemic, when cases are few in number, and hence sparsely distributed in the general population. In those cases, compartmental models may yield inaccurate estimates of likely spread.^7^

As well, if a model operates on the basis of a mass action incidence assumption (of homogeneous spread) then the model can only yield recommendations that relate to the population treated as a homogeneous entity. Where a service organization such as a regional health authority, or a regulatory/policy making body is setting out tactics for staged or localized restart of some but not all services, employing potentially different levels of protection in different settings, a model that works from the mass action assumption is inherently limited in its ability to project effects of different constellations of localized protections. If the model does not contain elements that relate to entities that are impacted by localized strategies or tactics to mitigate spread – or to reduce financial impacts of possible mitigations – then the model does not support decision-makers in rendering transparent the basis for measures that are being encouraged, promoted and possibly enforced.

Stated in slightly different terms – equation-based compartmental models, subject to mass action assumptions, are designed to reproduce historical data and predict future trends based on the systems of equations that describe those historical data. Agent-based models are designed to describe processes that take place within networks and give rise to those rates. This is illustrated graphically in the agent-based modeling stream of PHAC. ^8^ As such, these types of models can perform a distinctive function for decision-makers who have a delimited but direct span of control over networks of services associated with networks of movement of people through those services. An example would be a health region that delivers a broad array of secondary and tertiary services and must reconcile need for delivering health services with the attendant risk for healthcare associated transmission. An agent-based approach becomes particularly relevant when such a system is impacted by (and impacts on) local surrounding contexts in which risk is differentially impacted by selective/phased restart strategies that apply to various key entities (e.g., schools, restaurants/pubs, etc.) that cover the transmission topography.

### Motivation

CovidSIMVL is an agent-based model that can simulate transmission within a single bounded location (a “Universe”) or it can simulate transmission in a network of interacting universes (a “Multiverse” instance of a CovidSIMVL simulation). It is intended to achieve the following objectives:

1. <u>More accurate determination of the likely progress of epidemics within scenarios consisting of a delimited, heterogeneous array of Universes (a Multiverse).</u> This progression from one or a small number of infective persons to agents located in Universes within a Multiverse simulation is expressed in terms of the distribution of chains of different lengths that share common ancestors and the topological structure of the network of connections between chains. Constructs such as “superspreader events” would be expressed in terms of the topological features of transmission chains that emerge over iterations within CovidSIMVL trials.^9^ Different mixtures of chains, and the timing and localized contexts associated with the emergence of chains of different length carry different implications for risk mitigation.^10,11,12^
2. <u>More precisely timed and targeted testing in a network of interacting Universes, based on relative risk for transmission associated with component Universes, and based on lower/higher probability chains that span Universes within a Multiverse simulation</u>. This is accomplished in CovidSIMVL *via* the “Risk-per-Hour” (i.e., hours-to-next-infection) metrics. Universes with smaller values of this metric, or Universes that could achieve high degrees of adherence to protections that increase hours to next infection, would be those locations likely to benefit most from proactive testing and contact tracing
3. <u>Clearer understanding of the dynamics of spread within complex interacting systems.</u> The objective is to expose Universes (spaces) where most transmissions are taking place, and those which are least active in terms of transmissions.
4. <u>Generate sets of results associated with static configuration of key parameters, where variance reflects stochasticity at the level of movement and transmission within Universes</u>. Any observed outbreak, or any simulated outbreak incorporating elements of stochasticity, represents a single path profile for all possible paths conditioned by the pathophysiology of the infective organism and the local and population-based terrain that the outbreak traverses.^13^ The objective is to enable mathematical modeling of these results, thereby associating probabilities with outcomes associated with a series of trials employing the same parameters to depict scenarios. This is achieved *via* production of a version of CovidSIMVL that can be called as a function within R, in order to generate the requisite datasets and distributions.
5. <u>Provide a basis for generating estimates of the size of undetected fractions of infected populations based on distributions of transmission chains of different lengths in the population.</u> Better estimates of the total size of the infected population enables better estimates of key quantities such as Infected Fatality Rates.

## METHODS

### CovidSIMVL – Open-Source Agent-Based Modeling Tool

CovidSIMVL is a short-hand reference to “COVID-19 Simulation, Viral Load Version”. This is an agent-based infectious disease modeling tool that is configurable to generate products that characterize infection spread in local contexts.^14^

CovidSIMVL is an open source public domain system freely available under the GNU Open License Framework, and can be found at www.github.com/ecsendmail/MultiverseContagion. It is written in Javascript and runs in most modern browsers such as Chrome, Edge, Safari and Firefox.

The repository named above contains the Handbook in the directory /docs, which contains step by step instructions for running CovidSIMVL on the browser.

Different trials described in this report require different .csv files located in the above-mentioned github repository, in the /data directory:

- “population.csv”, “FiftyAgents.csv”, and “HundredAgents.csv” – to create populations of susceptible agents for trials
- “VL1.csv”, “VLfive.csv” – to insert different numbers of infective agents into trials.

These files need to be cloned or copied to the user’s local directory to run the simulations set out in this report.

Readers who wish to reproduce the trials set out in this paper should be aware that CovidSIMVL is an evolving system. For this reason, the README.md file should be reviewed before attempting to run the trials set out in this report. Note that the current version on github.com/ecsendmail/ MultiverseContagion has a parameter setting for MingleFactor (described later in this document) which differs from this paper by an order of magnitude. For example, what is shown in the Tables as 1.10 will need to be entered as 11 in newer versions of CovidSIMVL.

### Configuration of CovidSIMVL

#### Primary, Secondary, Tertiary Rules

CovidSIMVL enables model calibration by employing a hierarchically organized array of rules, to support the injection of biological, behavioural and local contextual heterogeneity/realism into models:

1. <u>Primary</u> – rules/parameters that embody physiologically ‘hard-coded’ features of the viral spread, such as usual incubation period or duration of infectivity.
  i. These rules may be modified to reflect differences in the response of individuals or clinical demographics to the infectious organisms, where those differences impact on key timings in the models. For example, if severity of illness is associated with viral load over time, primary rules can be adjusted to reflect different periods of time for key phases of viral transmission (incubation, infectiousness).
  ii. These rules are designed to capture the pathophysiologically necessary conditions for the possibility of infection.
2. <u>Secondary</u> – rules/parameters that determine whether/when a susceptible person becomes infected, or when an infectious person transmits to another person. For example, different degrees of physical proximity carry different risks for viral transmission.
  i. The CovidSIMVL ***HazardRadius*** and ***MingleFactor*** parameters function as a secondary rule that places risk for transmission within a spatial frame of reference.
  ii. By analogy, these secondary rules may be thought of as the proximal determinants (in both the causal and geospatial sense) of infectious disease transmission.
  iii. Secondary rules, together with primary rules, constitute necessary and sufficient conditions for the possibility of transmission.
  iv. Stochasticity enters into the simulations through these secondary rules and associated probability distributions to which they are keyed.
  v. However, these secondary rules do not determine the likelihood of transmission associated with movement of people ***between*** contexts. These are embodied in the Tertiary rules.
3. <u>Tertiary</u> – rules/parameters that prescribe patterns of behaviour for agents between local contexts.
  i. These bias odds for or against cross-over transmission from one context to another. As such, these rules determine dispersion of the infection beyond the boundaries of a given local context.
  ii. The CovidSIMVL ***schedules*** describe movements of agents between local Universes. They determine the manner in these local Universes interact to generate the dynamic of viral spread in a Multiverse model – see ***Appendix I***.
  iii. These tertiary rules may be thought of as the “distal determinants” of infectious disease transmission, in the sense that they condition the probabilities that transmission associated with the proximal determinants are actualized by a group bounded by a Multiverse scenario.

#### Setting population size to reflect saturation effects

The agent-based infectious disease simulation tool CovidSIMVL is based on a conceptual framework originally developed to describe biological systems, where two species interact in such a way that the actions of one group of agents (e.g., predators) results in a reduction in the size of another population or resource (e.g., prey). These types of models were generalized by Kermack & McKendrick^15^ in the 1920’s to provide mathematical descriptions of infectious disease transmission – where persons who carry an infection impact on populations of persons who are susceptible, in such a way that the number of infective or infected and recovered or deceased persons increases, while the number of susceptibles diminishes.

In the case of CovidSIMVL, this dynamic can be embodied in the generation of simulations where the number of susceptibles at the point of initialization is small enough, conditional upon settings for MingleFactor and HazardRadius, that saturation effects become important determinants of spread, or impacts of interventions that are introduced over time, e.g., vaccines.^16^ This is reflected in some of the simulations appearing in this report that self-extinguish.

#### CovidSIMVL Trials

Each set of parameters in CovidSIMVL represents a specific starting state. The stochastic nature of agent movements, viral transference, and initial spatial arrangement of the population makes each such trial unique, within confines set by the range of values that a given parameter can assume. The allowable ranges of values for parameters for a given series of trials locates a stochastically-varying set of results for those trials into a class that has statistical properties that characterize the likelihood of different outcome states.

#### Iterations within Trials

Each trial entails a number of iterations, each of which culminates in a description of number of key quantities, namely, the numbers of Susceptibles, Incubating Persons, Asymptomatic Infectives, Symptomatic Infectives, and Inerts (i.e., no longer impacted by close proximity to any other agents by virtue of having recovered or died or been effectively quarantined). Changes in those quantities over time reflect the dynamics of spread of the infectious organism and its carriers within the contexts built into the model.

The duration of the iterations can be set within a series of trials. For example, the duration could be set to an hour, which is useful when modeling dynamics that are associated with changes to agents within a 24 hour period, e.g., students spending a portion of the day in school, and a portion of the day at home.

A duration could be set for a day if the intention is to look at changes in rates over the course days, and there is no need or interest to factor in and portray changes at a more granular level

#### Stochasticity in CovidSIMVL, Reproducibility of CovidSIMVL trials

CovidSIMVL is a simulation program based on stochastic Monte-Carlo probabilistic generations of agent moves and viral dynamics in each generation. Each simulation is seeded with a base value for HazardRadius. However, for each generation (iteration) within a given run, this radius changes stochastically from a base value, according to a given agent’s viral load. As well, movement of agents within a local context over the course of generations is treated by CovidSIMVL in the MingleFactor as a random walk keyed to a Pareto distribution. The proportion of the local arena covered by random walks of the agents together determine the likelihood of contact between agents. The positioning of agents (also stochastic) determines the specific sequence of transmission from one agent to another.

Stochasticity in CovidSIMVL is located within changes to initial parameter values used to produce a simulated outbreak that unfolds over the course of several iterations. As such, multiple simulations for a given set of initial parameter values will not produce exactly the same results.

In order to glean information about the distributional characteristics of outcome measures (e.g., number of iterations to extinction of spread), multiple trials would need to be run. The calibration trials appearing in this report are intended to demonstrate that scalar values on key parameters *can* be set to values that reproduce a range of growth curves associated with different R_0_ values. Work is underway to determine what the underlying distributions look like for different sets of parameter values for populations of different sizes, moving between different Universes in a Multiverse simulation.

### Calibration

#### Approach

Calibration of CovidSIMVL occurs at three levels, reflecting the three levels of rules that together determine the outcome of a simulation, subject to the stochasticity built into the model.

1. Primary Rules – as detailed later in this document - these are calibrated to published information about differences in viral load associated with different phases in the course of the infection within an individual. Agents within/across Universes can be recalibrated should new data highlight different levels/periods of infectivity for different cohorts, e.g., children below the age of 10 in school, children above the age of 10 in school, etc.
2. Secondary rules – these rules operationalize a physical model used by CovidSIMVL to simulate spread of infection among agents. This physical model is embodied in two scalar quantities - HazardRadius and MingleFactor. The strategy for their calibration entails systematic variation of values on these two parameters to determine which combinations of values produce results that conform to the curves generated by equation-based models with the reproduction number R_0_ set to values <1, between 1 and 2, and greater than 2.
  a. HazardRadius – reflecting degree of infectiousness.
    i. This is a joint function of viral load, mechanism of transmission, e.g., fomite transmission, droplet (projectile) transmission, aerosol transmission, and physical proximity.
    ii. Physical proximity is a function of density of persons within a space and movement within the space.
    iii. HazardRadius can also be adjusted in a series of simulations, or midway through a set of iterations within a trial, to reflect different NPIs, such as physical distancing and/or wearing masks. For example, a lower value for HazardRadius could be set, either at initiation of a trial or midway through a simulated outbreak, to reflect a context where people are physically distanced, or functionally ‘distanced’ by wearing masks.
    iv. The HazardRadius is a single scalar quantity that can be adjusted in the simulation engine to reflect the joint contribution of these factors to the potential for viral transmission to occur.
  b. MingleFactor – this reflects movement within a space of a given size and density.
    i. It is set to reflect only movement, not the context within which the movement takes place. For example, MingleFactor for homeless persons in a congregate housing situation (e.g., an emergency shelter) would be set at a comparable level to persons in a pub. It would be set to a lower level for persons in a large grocery store, where density is lower.
    ii. MingleFactor is expressed as a single stochastically varying scalar quantity.
3. Tertiary rules – these are embodied in schedules that describe movement of agents over specified periods of time across component Universes that collectively constitute a Multiverse simulation.
  i. They are intended to reflect the movements and situations of people in the real world.
  ii. As well, Universes need to be seeded with initial values for numbers of infective persons. To the extent that these reflect real rates of infection over time within arrays of real-world contexts, e.g., children in elementary school settings, parents working in intensive care units, volunteers working in homeless shelters, the simulations will acquire more/less resemblance to real-world risk profiles.
  iii. In the Multiverse version of CovidSIMVL, it will be important to capture relative rates of real-world infection in multiple locations in order to generate realistic estimates of where greater/lower levels of risk are concentrated.

#### Calibrating HazardRadius and MingleFactor to R_0_

In standard epidemiological studies of contagions, R_0_ is the number of successful transmissions to susceptibles by infectious agents within the duration of infectivity of that agent.

R_0_ is estimated in equation-based modelling to reflect changes in the total number of identified cases within the population over time. Agent-based models work differently. They start with events associated with individual agents interacting with other individual agents. They count events and generate aggregate results from information that starts at the level of individuals who have properties that play out in one or more contexts.

CovidSIMVL, as an agent-based model of viral transmission, counts contacts and successful transmissions for each infective agent. The Multiverse version of CovidSIMVL counts chains of transmission associated with index cases. Within this agent-based paradigm, the reproduction number is seen as a property of the individual interacting in one or more contexts in a manner that engenders risk or protects from transmission. The model can supply a point in time R_0_ value associated with component locations or for a collection of locations that are bound together by people whose movements span the collection of contexts.

The calibration question that arises for CovidSIMVL can be expressed as follows: for what range of values do parameters such as MingleFactor and HazardRadius produce curves that have the same shape as compartmental model (equation-based) curves reflecting values of R_0_ falling in three ranges:

1. R_0_<1 – virus is extinguishing.
2. R_0_>2 – protective measures are failing to mitigate the inherent potential for each person to spread the infection to more than one person, given prevalent patterns of behaviour and the pathophysiology of viral transmission. This manifests as exponential growth.
3. R_0_>1 but < 2 – dynamic equilibrium when there remains a reservoir of susceptible persons interacting with infective persons, but transmission is being held in check by various mitigations such as social distancing, infrequent mingling, the wearing of masks, reducing time indoors, etc.

We have used the following parameters to run these trials:

1. The population size. Because CovidSIMVL operates within Universes of finite size, this population size is related to density.
2. The HazardRadius of the agents (uniform to start with at 2, 3, 4 and 5, and then varying stochastically).
3. The MingleFactor (degree of activity) of the agents: individually they are pre-set to 3, and then these are modified stochastically by a universal MingleFactor for the space, which modifies the activity of the individual as a product. That is, the final MingleFactor of an agent is the (individual MF) x (Universal MF).

#### Outcomes

For the material presented in the remaining portion of this document, we have run a series of trials with different parameter settings, to obtain two sets of outcomes:

1. For different combinations of HazardRadius and MingleFactor - the value of R_0_ at the time of the termination of a trial.
2. The values of the Critical Exposure Times in the deciles 1 to 5 to reflect rates of infection (detailed in Results section in the discussion of “Risk per Hour”).

## RESULTS

### Calibrating Primary Rules – temporal dynamics of infection within an agent

The Primary rules built into CovidSIMVL govern the temporal dynamics of emergence of infectivity within an agent. These temporal dynamics are keyed to changes in the viral load within a contacted/infective/recovered person over time. Timings for incubating/infective/recovered periods are based on a paper published by Xi, He et al.^17^, as shown in ***Figure*** 1.

**Figure 1.**
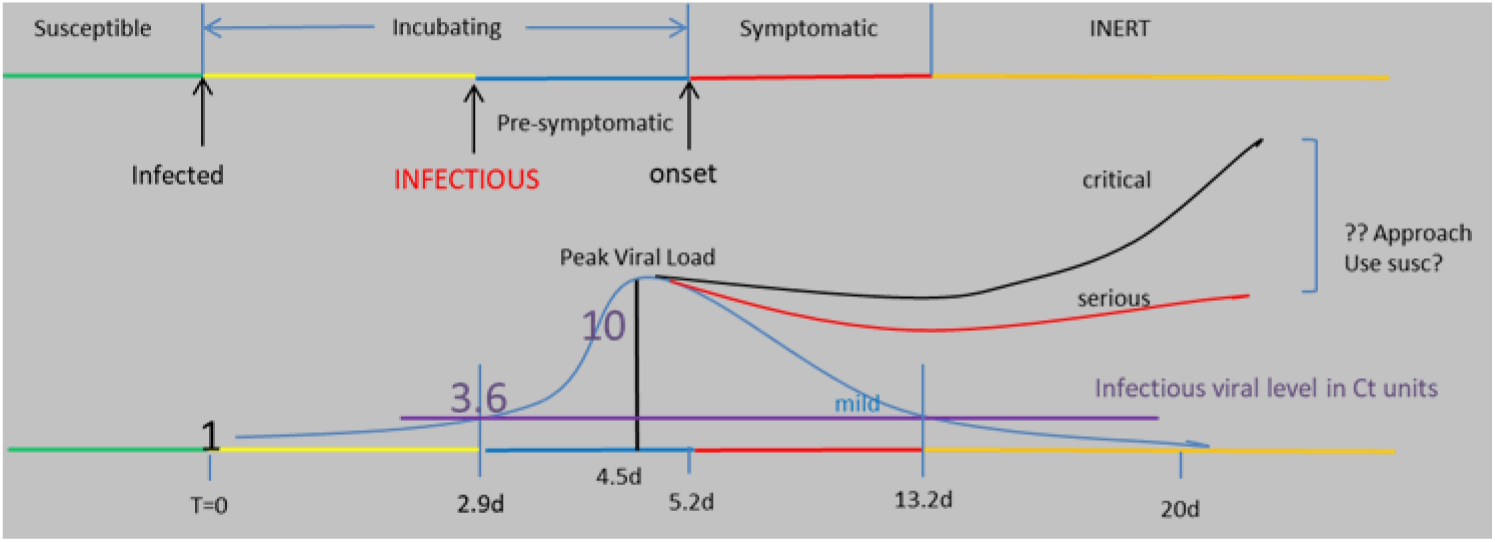
Configuring Primary Rules – Temporal Dynamics in Viral Shedding.

Temporal dynamics can be recalibrated should new data highlight different levels/periods of infectivity for different cohorts, e.g., children below the age of 10 in school, children above the age of 10 in school.

### Calibrating Secondar Rules – HazardRadius and MingleFactor

#### Sample CovidSIMVL trial

For the simulation depicted in Figure 2, the .csv files used are “2020.08.23HundredResOneU.csv” and “VL5.csv”, also found in the directory /data in the github repository. The Hazard Radius was set prior to the run (see Handbook) to 15, and the MingleFactor left unchanged at 10, with an initial set of five transmitters.

**Figure 2.**
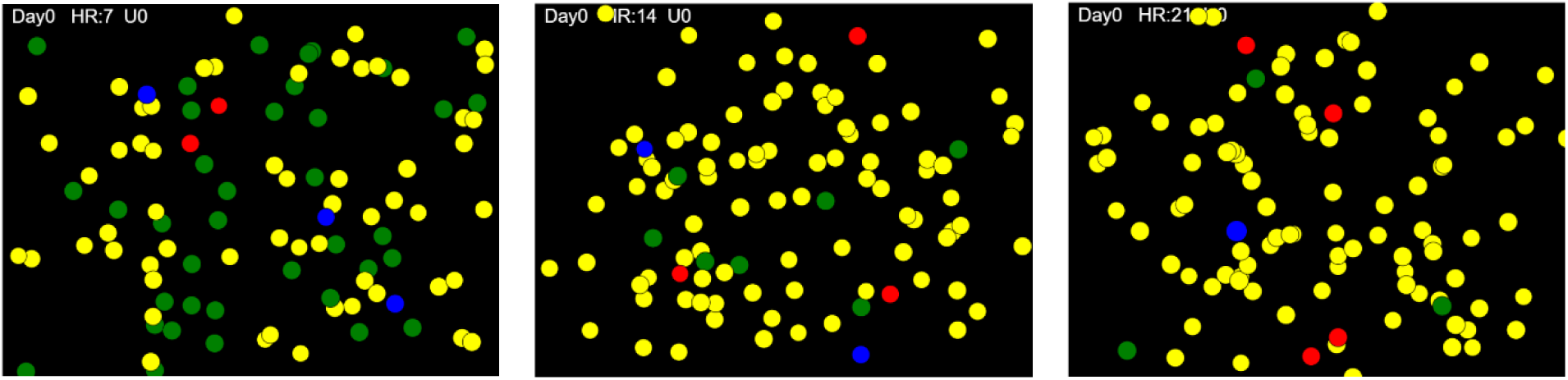
Single Universe Simulation, 63% infected after 7 iterations, 93% after 14 iterations, 97% after 21 iterations

#### Graphical conventions used to depict HazardRadius and MingleFactor

##### HazardRadius

because risk for transmission varies inversely as a function of distance, larger values of HazardRadius are used to characterize settings in which people are in closer physical proximity. In CovidSIMVL, this is reflected in the size of circles representing agents. See ***Figure 2***, below. The density of circles within the display reflects the distribution of agents within a field of finite dimensions.

##### MingleFactor

each iteration of CovidSIMVL produces a graphic depiction of the current state of the progress of an infection in one or more Universes. A series of such depictions of iterations within a single simulation captures the dynamic of spread. These dynamics are reflected in the distribution of agents in different stages of infectivity in ***Figure 2***. The uneven spacing reflects stochasticity of agent movement within the space, since the simulation is seeded with all agents evenly dispersed.

#### Graphical conventions used to depict Agent States in CovidSIMVL visualizations

Each agent can be in one of the following states at any given point in time:

Green = Susceptible

Yellow = Exposed and incubating

Blue = Aymptomatic or undetected but infective

Red = Symptomatic (detected) infective

Orange = Inert (e.g., recovered and assumed no longer infective; deceased; quarantined).

A HazardRadius of 15, together with a MingleFactor of 10, values will generally produce a rapidly spreading infection in CovidSIMVL that will affect all or almost all persons within a given space. For the example in the illustrative run depicted in ***Figure 2***, 63% of the 100 Susceptibles were infected after 7 iterations. Over the course of the next 7 iterations, the remaining number of Susceptibles had fallen to 7%. Over the course of the next 7 iterations, the number of Susceptibles was reduced to 3%. After 39 iterations 100% of the Susceptibles were infected. As will be shown in the following material, when different values are assigned to parameters, the simulated outbreak may self-extinguish before all Susceptibles have been infected.

#### Outcome Measures – Calibrating HazardRadius and MingleFactor to R_0_ Values

The R_<u>0</u>_ trials were performed by using a population of 100 identical agents, with a single initial transmitter, and Hazard Radius and MingleFactors set as per ***Table 1***. The trials were run either until 50% of the agents were still surviving (Green) or the trial self-extinguished. At that point, the R_0_ value was recorded.

**Table 1.**
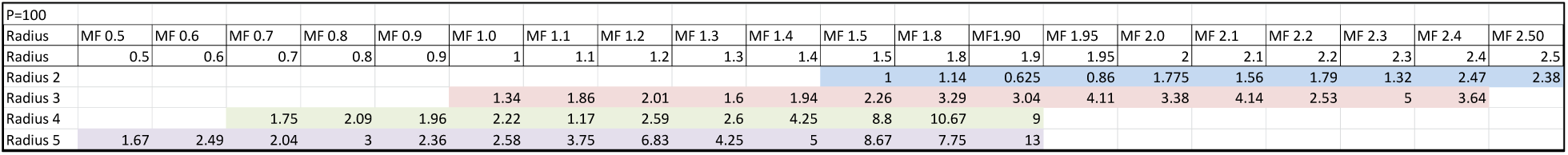
HazardRadius and MingleFactor for calibration trials

The CovidSIMVL simulation has the capacity to record every single transmission that an infective agent produces until the transmitted becomes inert (from Red symptomatic to Orange inert), about 10 days after onset of symptoms. Therefore at the end of the trial, the total number of transmissions by the Inerts, averaged, is the estimate of R_0_. The .csv files used for these trials were “HundredAgents.csv” and “VL1.csv” found in /data directory in the repository.

***Table 1*** shows a structured series of trials with variation in HazardRadius (Y axis) and MingleFactor (X axis). Cells contain R_0_ values at the end of the trials. Note in this table that there are specific ranges in which combinations of HazardRadius and MingleFactor produce epidemics that terminate with the population all infected, for which we report R_0_ values. For HazardRadius 4 and 5, we do not go beyond MingleFactor values that produce R_0_ > 10.

The corresponding graph for the trials as set out in ***Table 1*** is displayed in ***Figure 3***.

**Figure 3.**
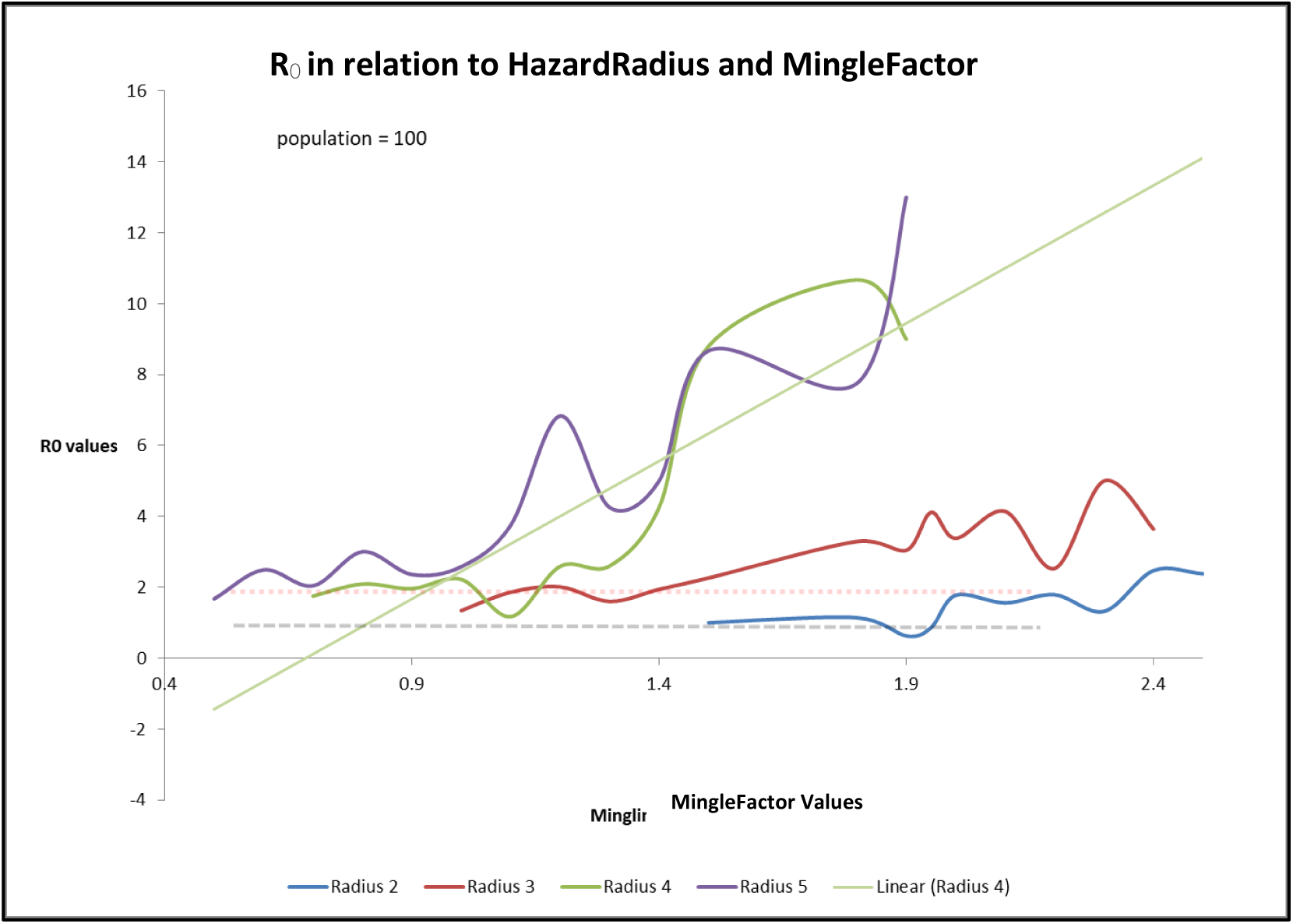
R_0_ in relation to HazardRadius and MingleFactor.

The important observations in ***Figure 3*** are:

1. For different HazardRadius values, the values of R_0_ falling between 1 and 2 (black and red dotted lines) are different for the distinct MingleFactors. For example, most values of R_0_ for HazardRadius of 5 are greater than 2 when MingleFactor is greater than 0.5.
2. For HazardRadius = 2, the values of R_0_ are consistently below 2, and for MingleFactor < 1.8, the values of R_0_ are < 1. Assigning these values to HazardRadius and MingleFactor results in trials which terminate before the target of 50% infected.
3. For each value of HazardRadius, the starting point at which R_0_ is meaningful is different. This is because, below these limits, the simulations terminate very early or do not proceed at all, that is, no contacts occur if the entities are very small and the movement is very small.

#### Estimates of Risk per Hour (“RPH”)

The results for calculating Risk per Hour and for the derivation of Transmission Trees are based on the ability of modern browsers to have a console.log, into which CovidSIMVL reported each transmission of the virus from one Named Agent to another Named Agent at the time (iteration) in which the transmission occurred. This time series forms the basis of the calculations of RPH and Transmission Trees.

A given trial may be set up to iterate until values are produced for a series of outcomes or threshold states:

1. The first 10% of agents newly infected
2. The point at which 20% of agents are infected
3. The point at which 30% of agents are infected
4. The point at which 40% of agents are infected
5. The point at which 50% of agents are infected

Note that for certain configurations of parameters, a particular trial may end before a threshold is reached. For example, the trial may end after only a few transmissions if there are no longer any infectious persons within the population specified for the trial. In other words, herd immunity is achieved in these trials before all persons have been infected.

We create the metric for the 10% threshold (“10% RPH”) as follows: we track the number of generations in the trial to reach 10% of the population as Newly Infected. For populations of 100, the number would be 10 new infections. If this took 100 generations, and the duration for each generation is 1 hour, then we have the following:

- The average number of generations for each newly infected = 100/10 = 10 generations.
- Within the first 10 new infections, the average span between infections is 10.
- The risk is 1/10 that there will be an infection in a generation in an hour in this time segment.
- The Risk per Hour (RPH) is defined as the inverse – i.e., 10.
- The larger the value for RPH, the lower the chance that there is an infection in an hour (a generation).
- The smaller the value for RPH, the more likely an infection will occur in the hour.
- With the progression of a trial, the RPH can be increasing or decreasing. This reflects a combination of an increase in the number of infectives and their dispersion within a finite space causing RPH to drop), together with saturation effects (causing RPH to rise).
- A falling RPH is an accelerating epidemic, while a rising RPH shows a decelerating epidemic.

The same procedure is employed to generate RPH metrics for 20%, 30%, 40% and 50% to capture the number of generations the trial required to reach those levels of infection.

Consider, for example, a trial that was set up to terminate as Newly Infected = 50%. At that point, the number of generations required to reach each interim/final goal state is recorded, as well as R_0_ at that point. The trials record appears in ***Table 2***. For the data in this table, the Hazard Radius was its default value of 5, and the same “HundredAgents.csv” and “VL1.csv” files were used, with MingleFactors set by the user at run time as shown. These data produced the graph in ***Figure 4***, although the data for MingleFactor 30 was not included in the ***Table2***.

**Table 2.** RPH values for different targets, holding HazardRadius constant and varying MingleFactors

| Population | HazardR | MingleF | 1st new | 10-Gen | 20-Gen | 30-Gen | 40-Gen | 50-Gen | 10%RPH | 20%RPH | 30%RPH | 40%RPH | 50%RPH | R0 |
| --- | --- | --- | --- | --- | --- | --- | --- | --- | --- | --- | --- | --- | --- | --- |
| 100 | 5 | 1.00 | 9 | 302 | 455 | 735 | 1065 | 1215 | 30.20 | 22.75 | 24.50 | 26.63 | 24.30 | 2.36 |
| 100 | 5 | 1.00 | 25 | 322 | 443 | 575 | 638 | 742 | 32.20 | 22.15 | 19.17 | 15.95 | 14.84 | 2.8 |
| 100 | 5 | 1.10 | 51 | 202 | 294 | 402 | 493 | 538 | 20.20 | 14.70 | 13.40 | 12.33 | 10.76 | 3.75 |
| 100 | 5 | 1.20 | 0 | 146 | 235 | 295 | 342 | 376 | 14.60 | 11.75 | 9.83 | 8.55 | 7.52 | 6.83 |
| 100 | 5 | 1.30 | 0 | 164 | 249 | 345 | 411 | 469 | 16.40 | 12.45 | 11.50 | 10.28 | 9.38 | 4.25 |
| 100 | 5 | 1.40 | 7 | 120 | 201 | 280 | 328 | 391 | 12.00 | 10.05 | 9.33 | 8.20 | 7.82 | 5 |

**Figure 4.**
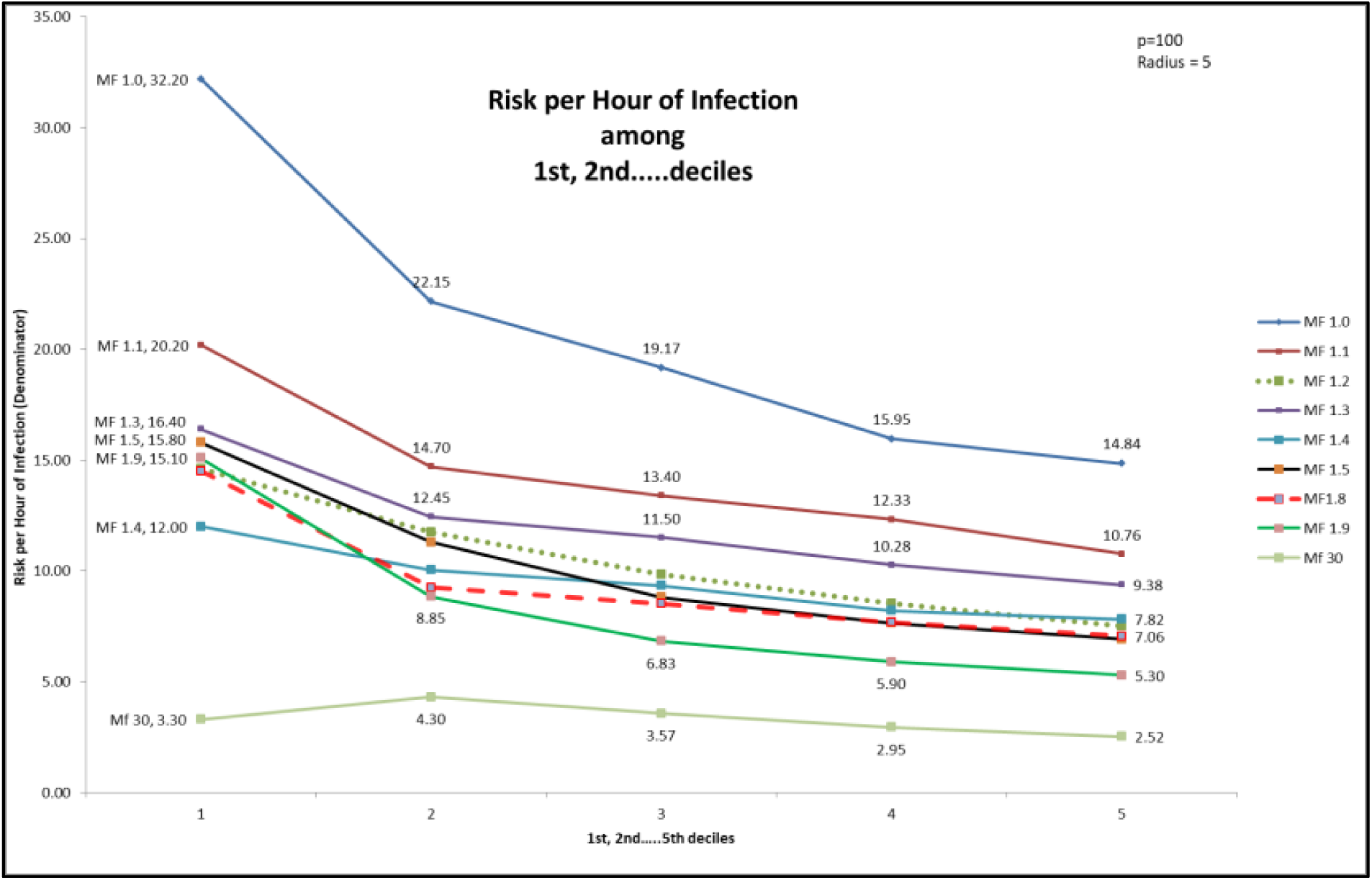
HazardRadius = 5, Rate per Hour among 1^st^…5^th^ deciles for varying MingleFactors

In ***Figure 4***, each line represents a different value of MingleFactor. Recalling that lower RPH values represent infections spreading at a higher rate per hour, the results appearing in this figure show that as MingleFactor rises, the epidemic progresses more rapidly as reflected in lower RPH values.

In this figure, the leftmost set of points, for the 10% decile, shows that a MingleFactor of 1.0 produced an RPH of 32.20, monotonically decreasing to and RPH of 3.30 (hours between infections as MingleFactor increases to 30.

For each line, as we advance in deciles along the trial, the values of RPH continue to fall. This indicates that the epidemic is accelerating (lower hours between infections) as it progresses. This is despite the removal of Susceptibles due to transmissions, which shows that as the epidemic progresses, the ratio of transmitters to Susceptibles continues to increase.

The dynamics depicted in ***Figure 4*** are keyed to a HazardRadius of 5 and a range of MingleFactors. Holding the range of MingleFactors constant but reducing the HazardRadius to 4 produces a different set of results, as depicted in ***Figure 5***.

**Figure 5.**
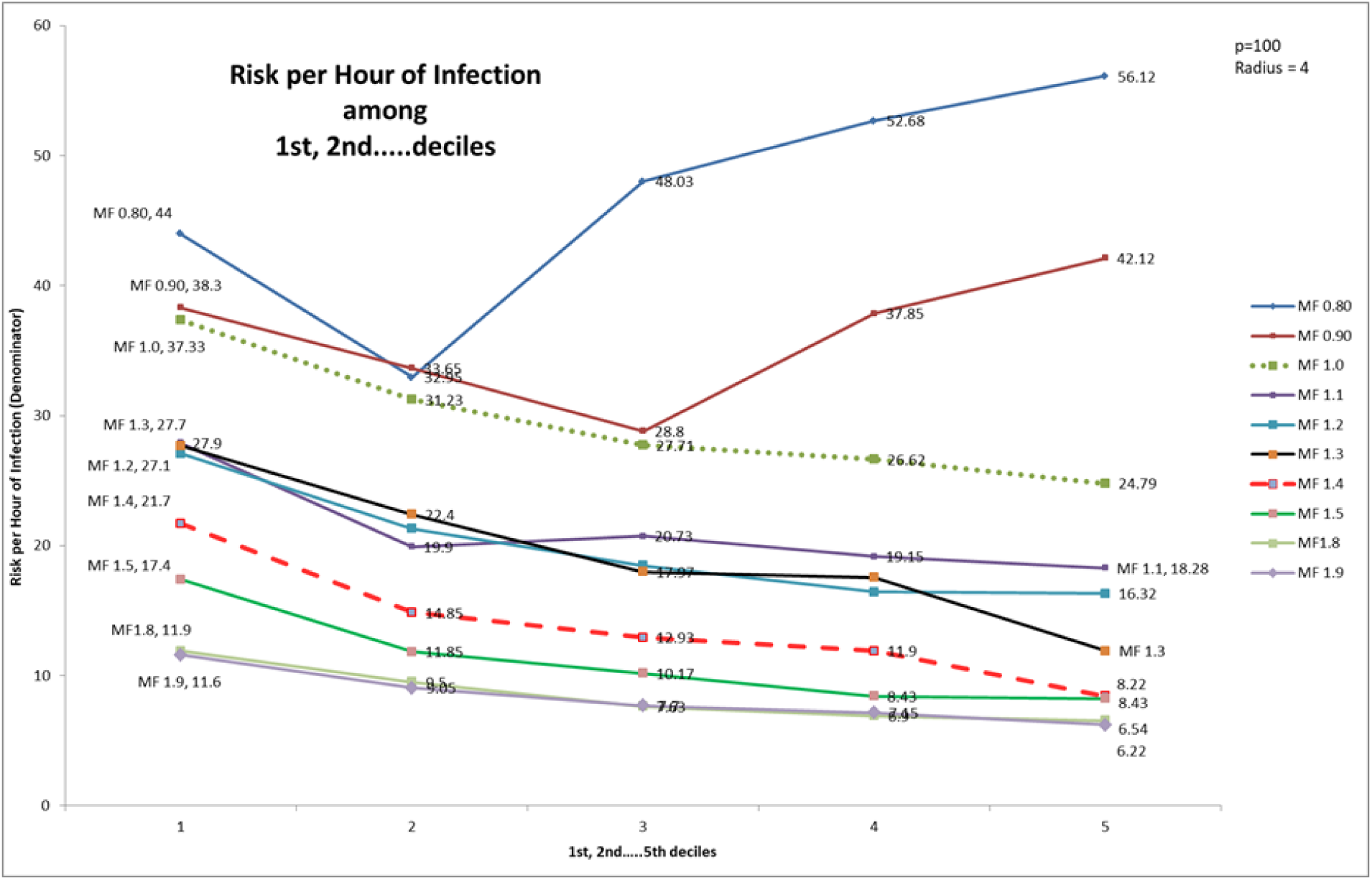
HazardRadius = 4, Risk per Hour among 1^st^…5^th^ deciles for varying MingleFactors

In this figure, the lines for MingleFactor 0.80 and 0.90 fall to the second decile and then have upward slopes. This indicates that the values of RPH are rising as the trials progress.

This rising RPH means that as the epidemic progresses, new infections occur more slowly. As such, this is the sign of an epidemic that is headed to self-extinction at best, and stability at worst.

High MingleFactors produce higher Risk per Hour metrics, because the agents are more mobile, and they are more likely to make contact in any particular generation within the model. The Susceptibles that are contacted and transform into infections diminish the total number of Susceptibles in the starting population and add to the number of infectives. This results in a greater effective density of infectives within the population, and greater risk for any of the uninfected (Susceptible) persons.

Where the decline is steep in Risk per Hour, we would expect R_0_ to be large, and where the Risk per Hour increases with deciles, we would expect R_0_ to remain within the range of 1-2, depending on number and effective proximity of available Susceptibles and the ‘effectiveness’ of protections as reflected in the parameters. In the case of a steep decline in Risk per Hour, the range for R_0_ values could drop below 1 if the rate of new infections per agent drops below a critical value related to their duration of infectiveness.

These trials have been set to run at hourly intervals for 24hrs per day. As such, with CovidSIMVL, we can model spread in which the same people have different transmission risk profiles depending on where they are located at different times during the day.

Take the example of a classroom, which has a student in it for 6 hrs a day. If the Risk per Hour is 30, or 1 in 30 hours, this might be optimistically interpreted to be 5 days at 6 hours/day with the risk being 1/30 per hour for an infection to take place. In 30 hours, we will get ONE infection, from this population.

By contrast, if the Risk per Hour for that student in a different setting is 6, or 1 in 6 hours, this might be interpreted to be 1 infection per 6 hours of classroom time (1 school day), with a risk per hour of 1/6.

## DISCUSSION

### Evaluating Policy Options via a Structured Series of Simulations

Various real-world policy options can be encoded via the parameters located within each of the three sets of rules that govern simulated dynamics in CovidSIMVL.

#### Primary Rules

reflecting within-agent viral growth dynamics – policies relating to testing could have the effect of shortening the infective period. This would be captured in the primary set of rules.

#### Secondary Rules

reflecting between-agent interactions in a fixed space – policies relating masks and physical distancing would be captured by HazardRadius and MingleFactor, the focus of this report on calibrating CovidSIMVL.

#### Tertiary Rules

reflecting the movement of populations between common spaces (Universes) – these are governed by schedules that reflect location/movement of agents between Universes over time. These schedules can be adapted to reflect different options re: selective/staged restart of various services, such as components of the health service system, places of work, education, or recreation.

These tertiary rules impact on chains of transmission – both their length for a given set of Universes assembled into a CovidSIMVL Multiverse, and the distribution of chains of different length. See ***Appendix I*** for a brief discussion of the Multiverse version of CovidSIMVL. See ***Appendix II*** for an initial exploration of chains of transmission generated by CovidSIMVL Multiverse simulations.

In keeping with the “all other things being equal” foundations of experimental scientific methods that can possibly yield interpretable results - agent-based tools such as CovidSIMVL can readily manage the configuration and implementation of a structured array of scenarios where select combinations of parameters can be varied, while others are held constant.

The CovidSIMVL trials reported in this document illustrate the correspondences between R_0_values falling within ranges that reflect distinct dynamics of spread and different configurations of HazardRadius and MingleFactor. These ranges supply a guide to setting parameters for modelling epidemics that display distinctive characteristics. As well, modifying the parameters with known characteristics to reflect changes in protections can, at a minimum, provide some insights into the possible impacts of those changes. Locating those changes within Universes in CovidSIMVL Multiverse simulations (see Appendix I) can provide insights into the possibility of optimizing impacts by localizing protections. Work based on the Multiverse version of CovidSIMVL is the subject of papers currently drafted that reflect work that is underway.

### Agent-Based Models, Equation-Based Models – ‘Validating’ CovidSIMVL – against what?

#### “There is no there there”^18^

Traditional compartmental models convey an understanding of epidemics by retrofitting equations to sets of datapoints that are treated as representative of “reality”. They are validated by demonstrating a close fit between real-world datapoints (e.g., number of test positive) and the curves generated by systems of equations. These curves have properties such as shape (e.g., rising, flat), elevation, and derivative properties such as rate of increase, summarized by metrics such as R0.

However, the ecological validity of a model that has been constructed to fit a context and time-bound set of datapoints is contingent on the validity of the assumptions (e.g., mass action incidence) in the model that enable its results to be extrapolated to other contexts – or even applied to the population on which the results have been derived, if that population does not in fact conform to the assumptions of the model. In particular, if the model assumes homogeneous spread among all susceptible persons in a population, the model may supply pertinent information if policies are going to be applied equally to all members of the population, regardless of local differences. However, if the assumptions are manifestly not true when the viral spread topography is viewed at a more granular/local level, the policies may not secure the scope and level of social acceptance necessary for them to produce the desired effect.

Heterogeneity of reported metrics such as R_0_ across geographic/political locale, as the SARS-CoV-2 pandemic unfolds, demonstrates that there is no one typical infection for a multi-host pathogen like SARS-Cov-2.^19^ As such, this report does not seek to validate CovidSIMVL by showing that it can reproduce an underlying reality associated with any particular set of real-world datapoints. Instead, it seeks to demonstrate *how* the rules can be configured in order to generate simulations of a variety of real-world scenarios that are characterized by a range of values on metrics such as R_0_. As demonstrated in this report, parameters in the single universe version of CovidSIMVL, which mirror that ‘standard’ tripartite set of parameters^20^ that govern spread in equation-based models (duration of contagiousness, likelihood of infection per contact, and contact rate) can generate results that reproduce the curves associated with different values of R_0_ that describe different real-world infectious disease dynamics.

#### What vs how?

CovidSIMVL does not seek to predict a real-world future state based on a set of historical data. This is a task for which equation-based approaches are more naturally suited – if the intent is to predict spread in large populations where heterogeneity is not pertinent to decisions supported by the predictions. It *does* seek to enable exploration of possible consequences of policies, implemented on a local level, in order to generate results that relate policies to change in total infections, or infections per hour, or length/distribution of chains of infections. In other words, the equation-based models may be suited to the task of predicting *what* might happen. CovidSIMVL is more focused on the question of *how* an infection such as SARS-CoV-2 could spread in an interacting set of local contexts in which a diverse array of factors can be varied systematically in order to highlight the relative impact of these factors in a population assumed to be heterogeneous in a variety of consequential ways.

## Data Availability

The manuscript contains results generated from an agent-based simulation modeling tool. The manuscript details key parameter values for the results generated with the CovidSIMVL tool. The tool itself is open-source and is available via Github at a location contained in the manuscript.

https://github.com/ecsendmail/AgentBasedModel

https://github.com/ecsendmail/MultiverseContagion

## APPENDIX I

### Multiverse Version of CovidSIMVL

#### Transmission within a Dynamically Interacting Array of Local Universes

CovidSIMVL is a simulation engine that models a *primary* set of rules (reflecting biologically-determined growth dynamics), and a *secondary* set of rules (between agent interactions in a fixed space – the subject of the above). These rules determine infection growth in a way that is intrinsic to the pathophysiological determinants of viral transmission (Primary rules), and to the interaction of those inherent characteristics of the virus with the physical proximity and movement of agents, regardless of the particular physical location in which they are moving (Secondary rules).

CovidSIMVL also generates simulations that reflect a ***tertiary*** set of rules. These are rules which enable the modeling of scenarios where individual agents spend time in different contexts (e.g., home, place of employment), and these different contexts may embody different levels of risk. For example, a person wearing PPE at work is less likely to become infected or transmit an infection than the same person participating in a group exercise class without a mask.

These tertiary level rules enable the depiction of scenarios that reflect transmission within an interlocking networks of component spaces (which we call Universes), such that

- Each Universe (e.g., homeless persons housed in a shelter) is relatively homogeneous with respect to risk for transmission by agents who are physically located within the particular Universe. However,
- The same agents in a different Universe (e.g., homeless persons presenting in an Emergency Department, or homeless persons entering into a withdrawal management program) may engage in different levels of risk-engendering or risk-mitigating behaviours that are characteristic of that particular location, and
- The same person may encounter different agents depending on location, e.g., a homeless person, and a collection of persons associated with an interlocking array of Universes may interact with different persons, depending on where they are located.

“School-based transmission” reflects a good example of interacting Universes coming together to form a single “Multiverse”. The infection may be contracted or transmitted to students/teachers in schools – and at least for high school or perhaps middle-school-aged children, a viral load that would be sufficient to transmit the infection to/from an adult would be sufficient to transmit the infection to/from another student, or teacher – or parent when the child returns home. Parents who work in healthcare could then transmit to/from other staff or patients in a healthcare location. In this scenario, school-aged children may transmit to parents but they will not transmit directly to patients. See ***Figure 6***, below for a screenshot of a multiverse configuration of CovidSIMVL that includes different locations with a school environment, home, a public recreational space such as a pub, and a place of work (long term care facility).

**Figure 6.**
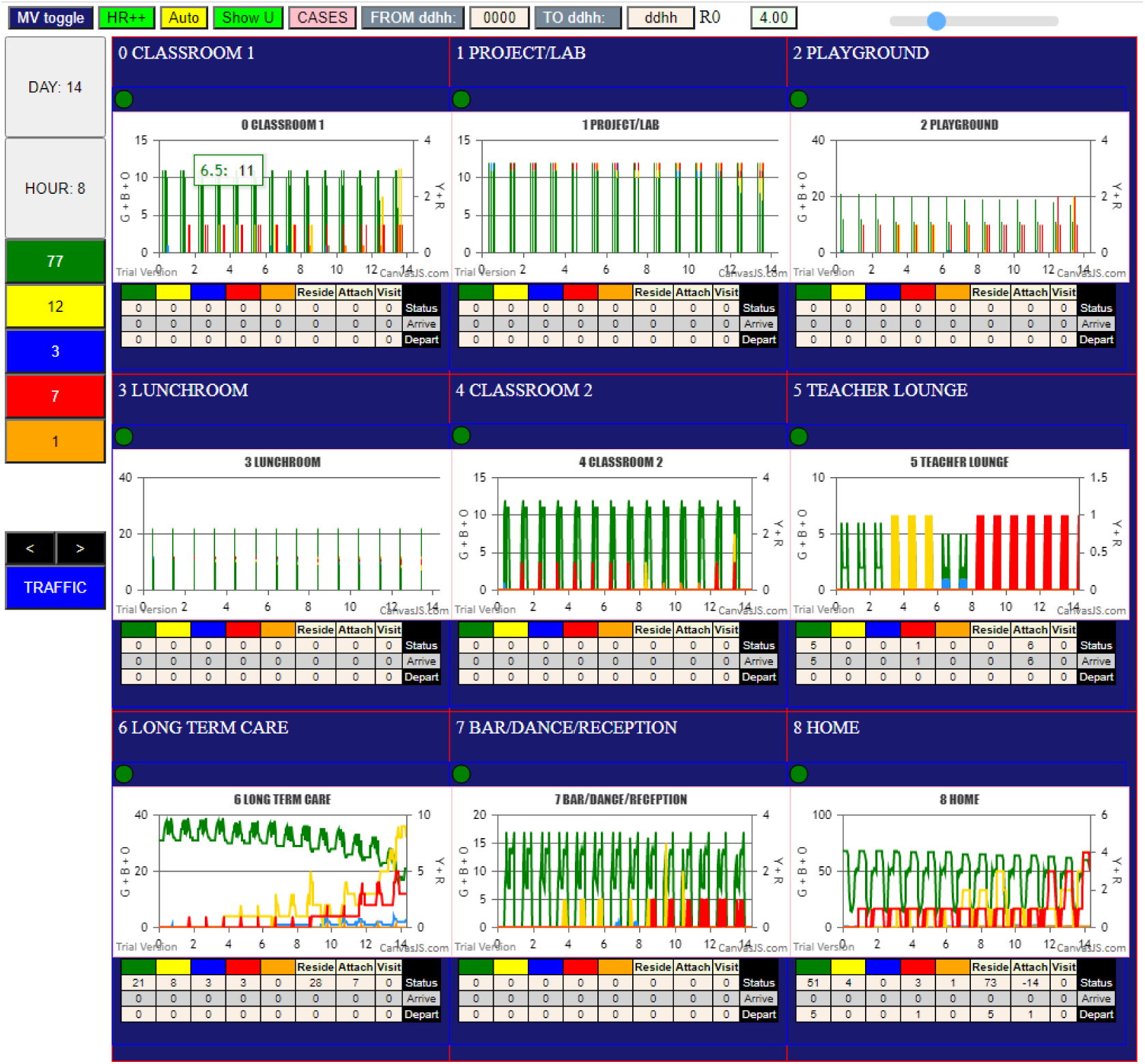
CovidSIMVL – Multiverse version, including schools homes, bar/pub, and a long-term care facility.

## APPENDIX II

### Transmission Chains

In the Multiverse version of CovidSIMVL, there can be chains of different length, spanning different component Universes, that link index cases to secondary or tertiary or n-ary cases.

The console.log in CovidSIMVL produces, infection by infection, a trace of who infected whom in what generation (time), in what Universe in a Multiverse simulation, identifying the family membership, and preceded by the sequence number of the new infection. The console.log looks like this:

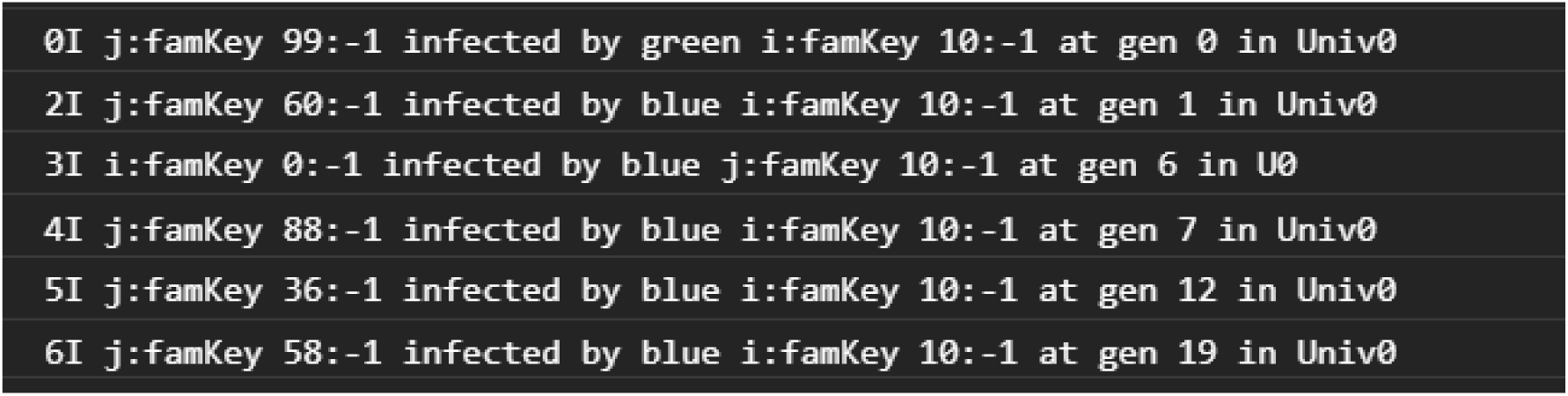

These data permit us to look more closely at the relationship between the length of the chains of transmission, and the frequency distribution of the numbers of infections that are incurred by infectious agents; in particular, whether there are super-spreaders as a side-effect of nature of contagion-based epidemics.

Here is a trace of infections with the parameters set with Population = 100, HazardRadius = 5, and MingleFactor for the Universe of 2, with the agent base MingleFactor set at 3, and the resulting R_0_ at 50 infected persons being 4.61, which indicates an expanding epidemic.

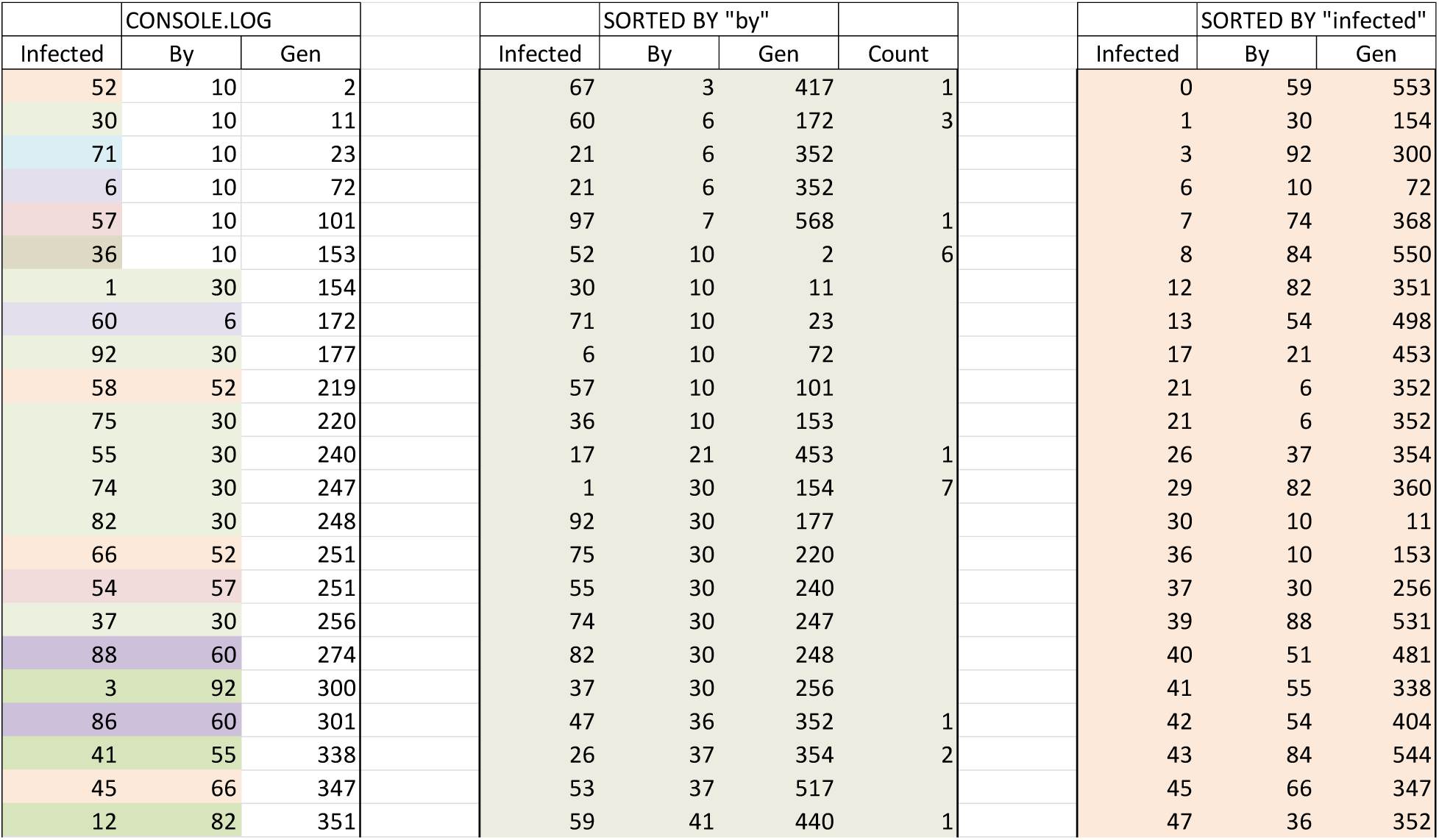
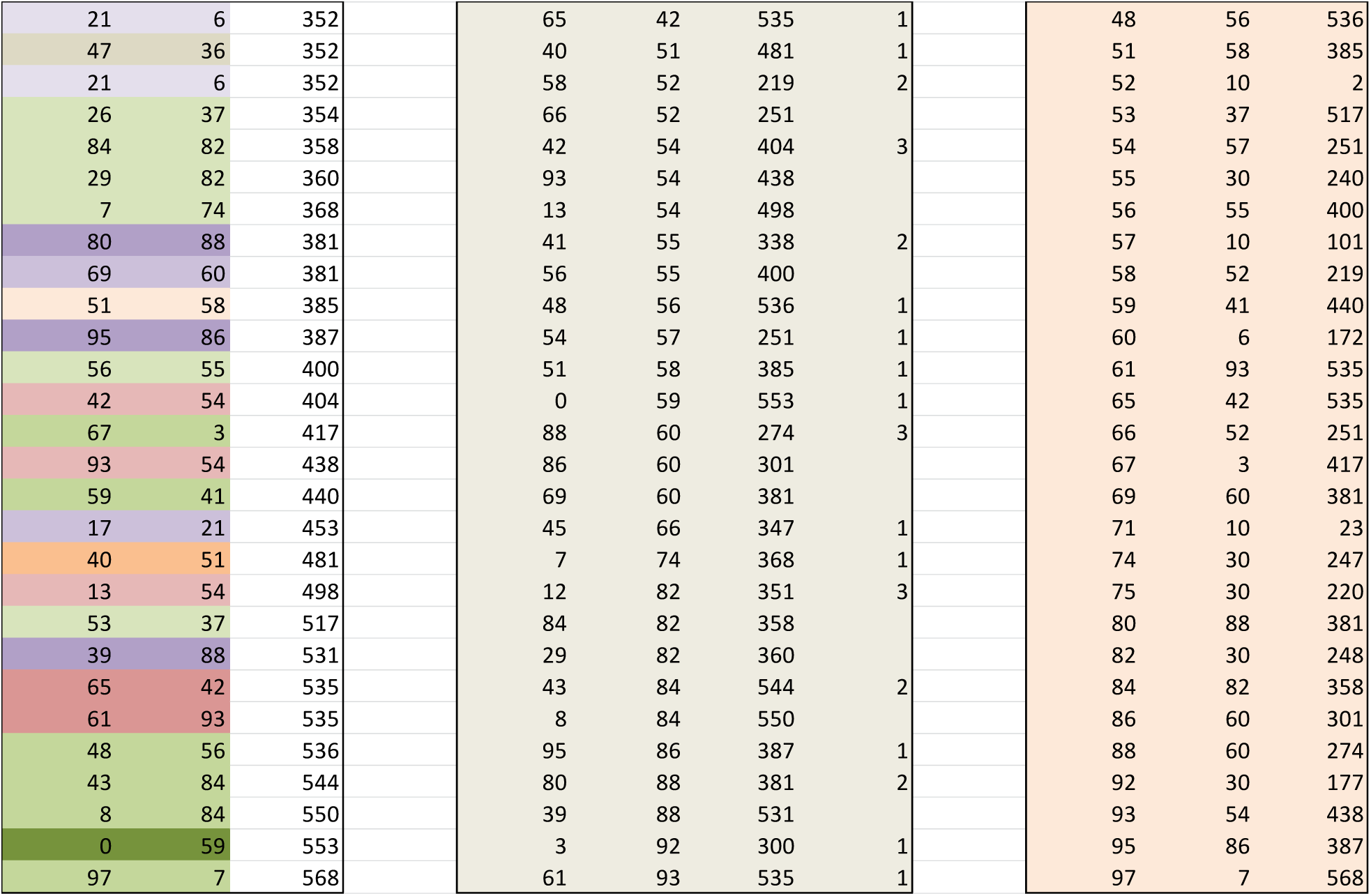

| CONSOLE.LOG |  |  | SORTED BY "by" |  |  |  | SORTED BY "infected" |  |  |
| --- | --- | --- | --- | --- | --- | --- | --- | --- | --- |
| Infected | By | Gen | Infected | By | Gen | Count | Infected | By | Gen |
| 52 | 10 | 2 | 67 | 3 | 417 | 1 | 0 | 59 | 553 |
| 30 | 10 | 11 | 60 | 6 | 172 | 3 | 1 | 30 | 154 |
| 71 | 10 | 23 | 21 | 6 | 352 |  | 3 | 92 | 300 |
| 6 | 10 | 72 | 21 | 6 | 352 |  | 6 | 10 | 72 |
| 57 | 10 | 101 | 97 | 7 | 568 | 1 | 7 | 74 | 368 |
| 36 | 10 | 153 | 52 | 10 | 2 | 6 | 8 | 84 | 550 |
| 1 | 30 | 154 | 30 | 10 | 11 |  | 12 | 82 | 351 |
| 60 | 6 | 172 | 71 | 10 | 23 |  | 13 | 54 | 498 |
| 92 | 30 | 177 | 6 | 10 | 72 |  | 17 | 21 | 453 |
| 58 | 52 | 219 | 57 | 10 | 101 |  | 21 | 6 | 352 |
| 75 | 30 | 220 | 36 | 10 | 153 |  | 21 | 6 | 352 |
| 55 | 30 | 240 | 17 | 21 | 453 | 1 | 26 | 37 | 354 |
| 74 | 30 | 247 | 1 | 30 | 154 | 7 | 29 | 82 | 360 |
| 82 | 30 | 248 | 92 | 30 | 177 |  | 30 | 10 | 11 |
| 66 | 52 | 251 | 75 | 30 | 220 |  | 36 | 10 | 153 |
| 54 | 57 | 251 | 55 | 30 | 240 |  | 37 | 30 | 256 |
| 37 | 30 | 256 | 74 | 30 | 247 |  | 39 | 88 | 531 |
| 88 | 60 | 274 | 82 | 30 | 248 |  | 40 | 51 | 481 |
| 3 | 92 | 300 | 37 | 30 | 256 |  | 41 | 55 | 338 |
| 86 | 60 | 301 | 47 | 36 | 352 | 1 | 42 | 54 | 404 |
| 41 | 55 | 338 | 26 | 37 | 354 | 2 | 43 | 84 | 544 |
| 45 | 66 | 347 | 53 | 37 | 517 |  | 45 | 66 | 347 |
| 12 | 82 | 351 | 59 | 41 | 440 | 1 | 47 | 36 | 352 |
| Infected | By | Gen | Infected | By | Gen | Count | Infected | By | Gen |
| 21 | 6 | 352 | 65 | 42 | 535 | 1 | 48 | 56 | 536 |
| 47 | 36 | 352 | 40 | 51 | 481 | 1 | 51 | 58 | 385 |
| 21 | 6 | 352 | 58 | 52 | 219 | 2 | 52 | 10 | 2 |
| 26 | 37 | 354 | 66 | 52 | 251 |  | 53 | 37 | 517 |
| 84 | 82 | 358 | 42 | 54 | 404 | 3 | 54 | 57 | 251 |
| 29 | 82 | 360 | 93 | 54 | 438 |  | 55 | 30 | 240 |
| 7 | 74 | 368 | 13 | 54 | 498 |  | 56 | 55 | 400 |
| 80 | 88 | 381 | 41 | 55 | 338 | 2 | 57 | 10 | 101 |
| 69 | 60 | 381 | 56 | 55 | 400 |  | 58 | 52 | 219 |
| 51 | 58 | 385 | 48 | 56 | 536 | 1 | 59 | 41 | 440 |
| 95 | 86 | 387 | 54 | 57 | 251 | 1 | 60 | 6 | 172 |
| 56 | 55 | 400 | 51 | 58 | 385 | 1 | 61 | 93 | 535 |
| 42 | 54 | 404 | 0 | 59 | 553 | 1 | 65 | 42 | 535 |
| 67 | 3 | 417 | 88 | 60 | 274 | 3 | 66 | 52 | 251 |
| 93 | 54 | 438 | 86 | 60 | 301 |  | 67 | 3 | 417 |
| 59 | 41 | 440 | 69 | 60 | 381 |  | 69 | 60 | 381 |
| 17 | 21 | 453 | 45 | 66 | 347 | 1 | 71 | 10 | 23 |
| 40 | 51 | 481 | 7 | 74 | 368 | 1 | 74 | 30 | 247 |
| 13 | 54 | 498 | 12 | 82 | 351 | 3 | 75 | 30 | 220 |
| 53 | 37 | 517 | 84 | 82 | 358 |  | 80 | 88 | 381 |
| 39 | 88 | 531 | 29 | 82 | 360 |  | 82 | 30 | 248 |
| 65 | 42 | 535 | 43 | 84 | 544 | 2 | 84 | 82 | 358 |
| 61 | 93 | 535 | 8 | 84 | 550 |  | 86 | 60 | 301 |
| 48 | 56 | 536 | 95 | 86 | 387 | 1 | 88 | 60 | 274 |
| 43 | 84 | 544 | 80 | 88 | 381 | 2 | 92 | 30 | 177 |
| 8 | 84 | 550 | 39 | 88 | 531 |  | 93 | 54 | 438 |
| 0 | 59 | 553 | 3 | 92 | 300 | 1 | 95 | 86 | 387 |
| 97 | 7 | 568 | 61 | 93 | 535 | 1 | 97 | 7 | 568 |

These numbers are derived directly from the console.log, using Excel to delete unneeded text, and separating the numbers with the Excel option in “DATA” of “Text to Columns”.

The first set of columns are by order of generations, with the Person Number of the Infected in the first column, the Infecting Person Number in the second column, and the third being the generation number. In some instances, the same infection is recorded twice, as in generation 351, Person 21, but it is only counted once.

The second set of columns duplicate the first set, but sorted using the “By” person number. This permits the easy tracking of a chain of transmission. For example, in the Generation set, the first entry shows Person 52 infected by Person 10. Going to the second set, and looking into the “By” column for 52, the entry shows that Person 66 was infected by Person 52, and we can continue this way.

The third set of columns is sorted by the “Infected” Person Number. This allows us to track the contagion in a backwards direction. For example, Person 95 in the first column, all the way down, was infected by Person 86, and in turn, locating 86, was infected by 60, and then finding 60, was infected by Person 6, and so on, finding 6 infected by 10, the Index case.

Using these data the longest chain was found to be:

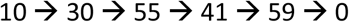

while the shortest was just

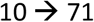

Tracking the tree of infections from the Index Case Person 10 through the 568 generations, we find that the distribution of Infectiveness is:

| Infects | Frequency |
| --- | --- |
| 1 | 16 |
| 2 | 5 |
| 3 | 4 |
| 4 | 0 |
| 5 | 0 |
| 6 | 1 |
| 7 | 1 |

The most frequent infectious intensity is one infection (16 of these events) while Person 30 infected 7 persons, and Person 10 (the Index Case) infected 6 persons. These do not tell us when these chains occurred.

The distribution of chain lengths (from the Index Case to a leaf in the transmission tree – either a node has become inert, or it has not infected any – had no descendants at the termination of the trial) are as follows for this trial:

| Chain Len | Number |
| --- | --- |
| 1 | 1 |
| 2 | 3 |
| 3 | 8 |
| 4 | 10 |
| 5 | 1 |

We can see that the most frequent length of transmission trains was 4, with 10 occurrences, and next were chains of length 3, with 8 such chains.

#### Chain Length – and the challenge of estimating the size of hidden populations

In expanding epidemics, the overall rate of infection is higher, so that one would expect shorter chains compared to epidemics that progress slowly or are self-extinguishing. Similarly, one would expect a higher average number of infections per infectious agent, with a more uniform distribution of infections per agent.

As infectiousness in CovidSIMVL is determined by HazardRadius and MingleFactor for defined population densities, it will be illuminating to identify the distributions of chain length and infectivity of agents over time, for varying parameters of HazardRadius and MingleFactors.

If these distributions can be characterized, it may then be possible to take tracking data from the field as samples into these distributions, and use the distribution of the chain lengths of the samples, fit them to the distributions from the synthetic epidemics, in order to estimate the hidden true population of infected.

